# Exploring Causal Association between Neuroticism Subclusters and Risk of Coronary Atherosclerosis: A Mendelian Randomization Analysis based on Genome-Wide Association Studies

**DOI:** 10.1101/2025.02.19.25322578

**Authors:** Jie Ming, Rui-lin Li, Yi-li Gao, Shu-ting Huang, Xiao-yan Yao, Qi Tan, Ming Zong, Lie-ying Fan

## Abstract

**Background:** Understanding the risk factors associated with coronary atherosclerosis (CA)—an evident cardiovascular disease—is vital. This study aimed to explore the causal relationship between neuroticism subclusters and CA, focusing on lipids as mediators, using genome-wide association studies and Mendelian randomization (MR).

**Methods and Results:** Instrumental variables were selected based on their genetic associations with neuroticism subclusters and CA. Causal effects were primarily assessed using inverse variance-weighted (IVW) methods. Sensitivity analyses and colocalization analyses were conducted, and possible confounders were determined using multivariable MR analysis. IVW modeling revealed that neuroticism subclusters significantly affect CA risk (*ebi-a-GCST006475* odds ratio [OR]: 1.02, 95% confidence interval [95%CI]: 1.01–1.03, *P* = 1.52E-05; *ebi-a-GCST006478* OR: 1.01, 95%CI: 1.00–1.02, *P* = 0.026; *ebi-a-GCST006950* OR: 1.01, 95%CI: 1.00–1.02, *P* = 0.036). Cochran’s Q test demonstrated moderate heterogeneity (*ebi-a-GCST006475*, *P* < 0.05, *I²* = 28%). The Steiger test confirmed the correct causal direction (*P* < 0.05). Multivariable MR remained significant after adjusting for confounders (*P* < 0.05). Mediation analyses suggested that lipids partially mediated the neuroticism–CA effect.

**Conclusions:** Overall, our findings indicated a causal link between neuroticism subclusters and CA, with lipids partially mediating this relationship, emphasizing the role of psychosocial factors in cardiovascular risk assessment and intervention strategies.

## INTRODUCTION

Cardiovascular diseases (CVDs) remain the leading cause of mortality worldwide, accounting for 31% of all deaths and approximately 18 million fatalities annually ^1^. Coronary atherosclerosis (CA) stands out as the primary cause of death among CVDs, accounting for nearly 45% of all CVD-related fatalities, highlighting its significant threat to global health ^2^. CA is a vascular disorder characterized by inflammation of the arterial walls, resulting in the accumulation of low-density lipoprotein cholesterol (LDL-C), monocytes, macrophages, and foam cells at inflammation sites ^3^. This gradual accumulation of plaque narrows the arterial lumen and reduces their elasticity, thus hindering the blood flow and resulting in serious conditions, such as coronary artery blockage, angina, or heart attack ^4^. The management of CA exhibits certain major challenges, including the inability to completely reverse the disease and stop its progression despite advancements in medical therapies and surgical interventions. Additionally, current diagnostic techniques such as coronary angiography and coronary computed tomography angiography associated with inherent risks are expensive. These issues underscore the urgent need for further research on the fundamental mechanisms of CA and the discovery of new therapeutic targets.

Neuroticism—recognized as one of the big five personality traits—is characterized by a tendency to experience various unpleasant feelings such as depression, anxiety, and irritability ^5^. Importantly, neuroticism is significantly heritable ^6^, with approximately 48% of the phenotypic variance attributed to genetic influences in twin and family studies ^7^. Recent genome-wide association studies (GWAS) have uncovered 136 distinct genome-wide significant loci linked to neuroticism ^8–11^, accounting for approximately 4.2% of the variation in neuroticism, with considerable polygenic overlap between neuroticism and diverse CVD risk factors ^12^. Furthermore, mental health disorders contribute to CVD onset ^13^, and an association of elevated neuroticism has been reported with diminished mental and physical health ^14^ and with an increased risk of all-cause mortality ^15^. Moreover, certain subclusters of neuroticism, such as depression and worry, serve as independent risk factors for CVD, including CA, indicating a robust association ^16^. Additionally, two other studies consistently documented elevated levels of inflammatory markers, such as interleukins 1, 6, and 17, in individuals with neuroticism ^17,18^. Therefore, neuroticism may trigger chronic inflammation damaging the vascular endothelium, which is an important factor in CVD progression ^19^. In addition to personality traits, dyslipidemia—characterized by elevated triglycerides (TG), LDL-C, and apolipoprotein B (ApoB) levels and diminished high-density lipoprotein cholesterol (HDL-C) and apolipoprotein A (ApoA) levels—also contributes to and is an important risk factor for CA ^20,21^. Furthermore, dyslipidemia is also associated with lifestyle factors, such as changes in physical activity levels and dietary intake patterns ^22–24^. Notably, personality traits affect lifestyle, cognition, motivation, and behavior ^25^. Previous studies have reported associations of optimism with high HDL-C and low TG levels ^26^, depression with low HDL-C and high TG levels ^27^, and total scores for anxiety, worry, depression, and other moods with high blood lipid levels ^28^. However, the causal associations among neuroticism, lipids, and CA are not yet fully understood, warranting further research. Despite studies suggesting a correlation between neuroticism and CVD, reports of increased risks of CVD and mortality in individuals with increased neuroticism remain inconsistent, and the link between the two has not been clearly established ^29–31^. The challenge of establishing causality can be attributed to the potential for reverse causality and unmeasured confounders in prospective clinical and epidemiological studies. Furthermore, randomized controlled trials (RCTs) with substantial sample sizes investigating the causal relationship between these diseases remain limited.

Mendelian randomization (MR)—an epidemiological method used to infer the cause and effect of a disease—can solve this issue to a certain extent using Mendel’s second law of inheritance to circumvent the bias of observational studies by treating genetic variations as instrumental variables (IVs) ^32^ and randomly assigning alleles, similar to the randomization process in RCTs. The genotype that determines the phenotype associated with the disease is not affected by environmental or confounding factors after the disease onset. Therefore, MR provides an alternative method for exploring causality with slight or no reverse causality or confounders ^33,34^.

Therefore, this study aimed to clarify the causal relationship between neuroticism subclusters and CA risk, emphasizing the mediating influence of lipid levels, thus offering new insights for CA prevention and management.

## METHODS

### Research Guidelines and Study Design

This study employed two-sample MR (2SMR) and an accessible dataset to investigate the causal relationship between neuroticism subclusters and CA risk as well as the mediating influence of lipids on CA progression among neuroticism subclusters. The study was conducted in accordance with the MR guidelines for strengthening the reporting of observational epidemiological studies using MR (the STROBE-MR Statement) ^35^. The research framework is presented schematically in Figure 1.

**Figure 1.**
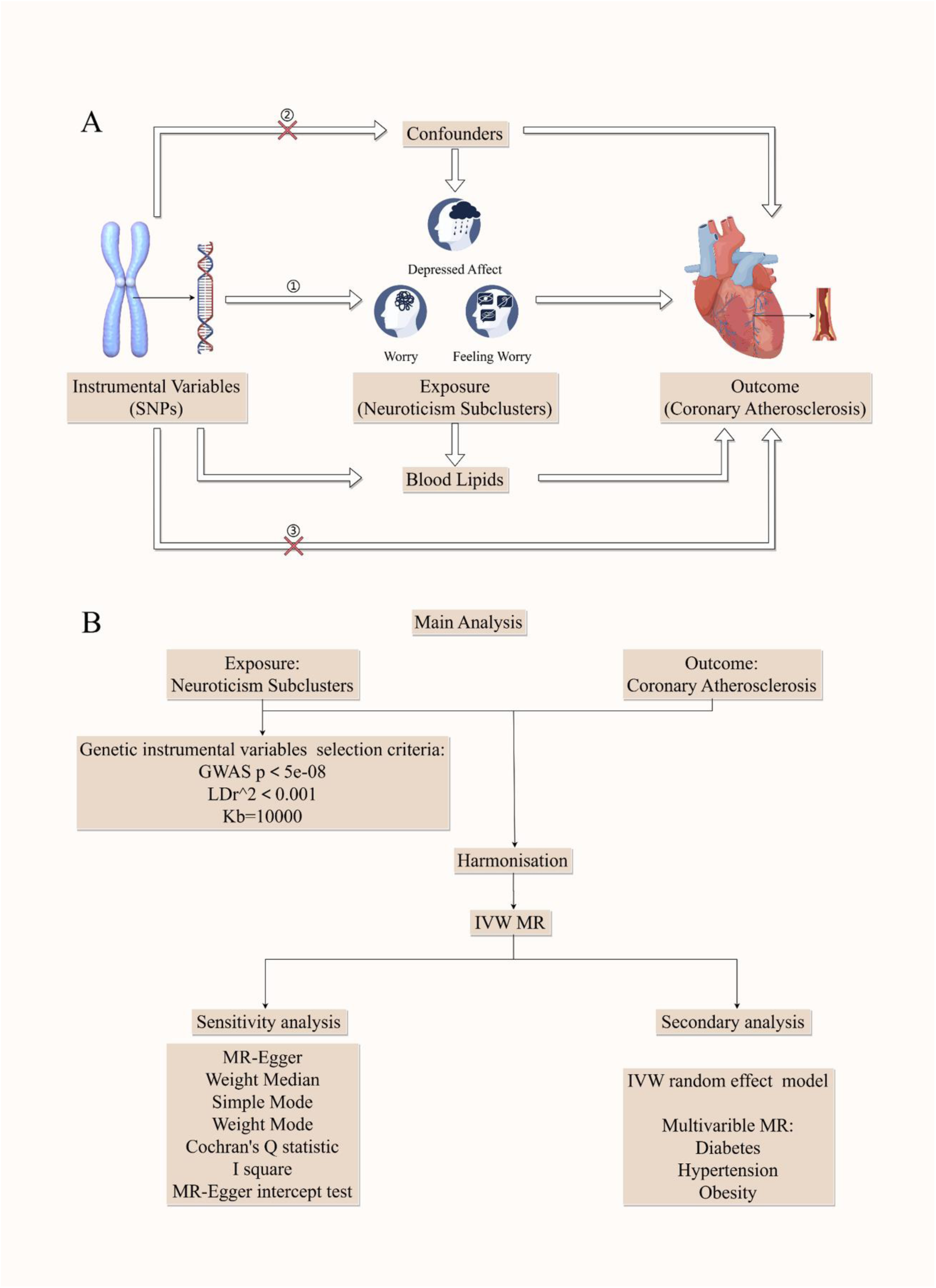
Technology Roadmap. A. The basic assumptions of MR analysis, including (1) the association assumption, which stipulates that the selected IVs must demonstrate a significant correlation with the exposure variable; (2) the independence assumption, ensuring that the IVs do not correlate significantly with potential confounders that may affect either exposure or outcome; and (3) exclusivity, whereby the IVs influence the outcome solely through the pathway defined as ’IVs → exposure → outcome’. B. Flow chart of analysis methods in this study. SNP: single nucleotide polymorphism; IVW: inverse variance weighted; MR: Mendelian randomization; GWAS: genome-wide association study; LD: linkage disequilibrium; MR-Egger: Mendelian randomization-Egger.

### Data Sources

Initially, we obtained the complete GWAS data from a publicly accessible database available at IEU OpenGWAS project ^36^. A list of neuroticism subclusters (Depressed affect: *ebi-a-GCST006475*, Worry: *ebi-a-GCST006478*, Feeling worry: *ebi-a-GCST006950*) based on existing literature was then compiled ^37^, and the respective GWAS data pertinent to these subclusters were incorporated into the downloaded dataset. We selected samples of European descent for subsequent analyses.

Regarding CA, we sourced the GWAS data (*ukb-d-I9_CORATHER*) from the previously acquired dataset, which exclusively comprised European samples. Additionally, we reviewed the literature to identify relevant risk factors for CA ^38^, integrating the corresponding GWAS data from the downloaded dataset: obesity (*finn-b-E4_OBESITY*), diabetes mellitus (*ukb-a-306*), and hypertension (*finn-b-I9_HYPTENS*). All GWAS data utilized were derived from samples of European ethnicity.

A list of lipid components (such as TG, HDL-C, LDL-C, Apo B, and Apo A) was prepared from the existing literature ^39^. The GWAS data associated with lipid levels were included in our downloaded dataset, ensuring that samples from European populations were retained for further analysis.

### IV Selection and Intensity Assessment

Three fundamental assumptions ^32^ must be satisfied for genetic variants to serve as valid IVs. First, the association assumption, which stipulates that the selected IVs must demonstrate a significant correlation with the exposure variable. Second, the independence assumption, which ensures that the IVs do not correlate significantly with confounders that might influence either the exposure or outcome. Third, the presumption of exclusivity, in which the independent variables affect the result exclusively through the pathway delineated as “IVs → exposure → outcome.”

We also established the criteria for identifying single nucleotide polymorphisms (SNPs) as IVs. SNPs from the exposure GWAS were selected based on a significance threshold of *P* < 5 × 10^-8^. SNPs exhibiting linkage disequilibrium, defined as r² < 0.001 and a physical distance exceeding 10,000 kb between genes, were omitted. Thereafter, GWAS data pertaining to the outcomes were retrieved based on the selected SNPs. Additionally, the *F* statistic was computed to evaluate potential weak instrument bias, wherein an *F* statistic value of <10 indicates a weak genetic variant, which might introduce bias into the findings ^40^. Consequently, such variants were omitted to preserve the integrity of the results. The *F* statistic is calculated as follows:

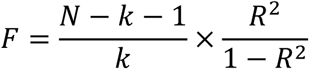

where N is the sample size, k is the number of IVs used, and R^2^ reflects the extent to which the IVs explain the exposure.

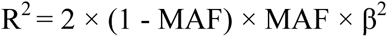

where MAF is the minimum allele frequency, and β is the allele effect value.

### Estimation of Causal Effects via MR

Multiple MR methodologies were employed to assess the causal relationships between neuroticism subclusters and CA, including the inverse variance-weighted (IVW), Mendelian randomization-Egger (MR-Egger), weighted median (WM), and simple and weighted modes. We mainly focused on the IVW approach, which is based on Wald estimations derived from meta-analyses of each SNP included in the analysis to derive the overall effect of exposure on the outcome. Additionally, we utilized the WM and MR-Egger methods as supplementary tests for MR estimation. The WM approach is particularly advantageous in scenarios where half of the genetic variation is deemed invalid ^41^. Furthermore, the MR-Egger intercept was used to assess the robustness of the IVs in relation to the direct-effect hypothesis, which asserts that pleiotropy is independent of the association between the variable and exposure ^42^, with *P* < 0.05, indicating pleiotropy.

The IVW method, which employs regressions that omit an intercept term and uses the inverse of the outcome variance as a weight ^43^, is more effective than alternative methods under certain conditions. Consequently, we implemented the IVW method as the principal analytical approach for MR analyses in the absence of pleiotropy, whereas the remaining four methods were employed as supplementary analyses. Additionally, the IVW random effects model was applied in cases of heterogeneity ^43^, and results were computed using the MR-Egger method if pleiotropy was detected. Finally, we performed a Steiger directionality assessment using the 2SMR package to examine the directionality of causation. In addition, we investigated the reverse causal effects to ascertain whether the outcome could potentially affect exposure using the same methodology.

### Sensitivity Analysis

Sensitivity analyses were performed to verify the robustness of our findings using various methods, including tests for heterogeneity, pleiotropy, and leave-one-out tests. The Cochran’s Q test was used to assess heterogeneity among individual SNP estimates. A statistically significant result indicated the existence of variability in the analysis. Notably, although Cochran’s Q test determines the existence of heterogeneity, it does not provide insights into its distribution. Therefore, the I^2^ statistic was used to reflect the proportion of the heterogeneity of the IVs in the entire variation. I^2^ ≤ 0, 0 < I^2^ ≤ 25%, 25% < I^2^ ≤ 50%, and I^2^ > 50% indicated no, mild, moderate and high heterogeneity, respectively. The precise calculation formula is as follows:

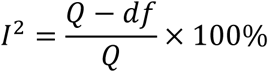

We performed an MR-Egger analysis to evaluate pleiotropy among the IVs. *P* < 0.05 for the MR-Egger intercept indicated significant pleiotropy in genetic variance. We then calculated the MR results, excluding individual SNPs, to assess their influence on the relationship between the neuroticism subclusters and CA. If the difference between the MR effect and aggregate effect estimate after excluding an IV was large, then the MR effect estimate was considered sensitive to the SNP.

### Colocalization Analysis

Colocalization analysis is used to investigate whether the association between different genetic variants (e.g., SNPs) and different phenotypic traits (e.g., disease or physiological traits) is driven by the same causal variant in a given region, thereby strengthening the evidence for an association between the two phenotypes ^44^. The phenotypes under consideration encompass both molecular characteristics and prevalent dichotomous diseases. Colocalization analysis evaluates whether identical genetic variants are linked to different phenotypic attributes and specifically assesses whether both phenotypes have a shared causal variant within a designated genomic region ^45^. Colocalization analyses were performed using the R package coloc, applying the Bayesian methodology ^46^ to test the following five hypotheses: H0: no significant correlation exists between phenotypes 1 (GWAS) and 2 (GWAS) with any SNP loci within the defined genomic region; H1/H2: phenotypes 1 and 2 do not exhibit a significant correlation with any SNP locus within the specified genomic region; H3: phenotypes 1 and 2 are significantly associated with SNP loci in defined genomic regions but are driven by distinct causal variant loci; and H4: phenotypes 1 and 2 exhibit significant associations with SNP loci in a specific genomic region and are influenced by the same causal variant loci.

To estimate the posterior probability of shared variants for colocalization among GWAS studies, we recognized the SNP with the lowest *P*-value from the exposed GWAS association as the lead SNP. Subsequently, we compiled all SNPs located 50 kb upstream and downstream of the lead SNP for colocalization analyses. We considered *P*H4 >0.75 and >0.5 as evidence of high and moderate colocalization, respectively.

### Multivariable MR (MVMR) and Estimation of Mediated Effects

MVMR extends traditional MR by employing genetic variations associated with multiple relevant exposures to ascertain their collective influence on a single outcome ^47^ and also examines the direct effects of each exposure on that outcome ^48^. Two-step MVMR can be used in mediation analyses to decompose the impact of an exposure on an outcome, estimating the causal relationships among the three variables through the mediating variable^49^.

We analyzed multiple relevant risk factors for CA (*finn-b-E4_OBESITY*, *ukb-a-306*, *finn-b-I9_HYPTENS*). Furthermore, the direct effect of the neuroticism subclusters on CA was determined via MVMR (case-by-case) using the neuroticism subclusters (*ebi-a-GCST006478*, *ebi-a-GCST006475*, *ebi-a-GCST006950*) obtained after screening.

We could obtain the indirect effect of neuroticism subclusters → lipids → CA pathway by the effect of neuroticism subclusters on lipids obtained after screening (Fig. 1). The value and standard error of the mediated effect were calculated using the following equations:

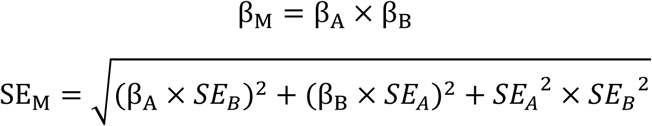

where β_M_ is the effect value of the mediating effect, β_A_ is the MR effect value of the neuroticism subclusters on lipids, β_B_ is the direct effect value of lipids on CA, SE_M_ is the standard error of the mediating effect, SE_A_ is the standard error of the MR analysis of the neuroticism subclusters on lipids, and SE_B_ is the standard error of the MR analysis of lipids on CA.

Combined with the causal stepwise regression method, if both β_A_ and β_B_ were significant, the indirect effect was significant, whereas if one of β_A_ and β_B_ was not significant, the Sobel test was used to determine whether β_M_ was significant or not, and if β_M_ was significant, the indirect effect was significant. In cases when the indirect effect was significant, if β_M_ was heteroscedastic with the MR effect value β_C_’ of the neuroticism subclusters on CA in the multivariate MR, the proportion of the direct effect accounted for by the neuroticism subclusters mediation of CA via lipids was calculated as |β_M_/β_C_’| × 100%. Furthermore, if β_M_ was heteroscedastic with the MR effect value β_C_’ of the neurotic subcluster on CA in multivariate MR, β_C_’ is the same sign, then the proportion of the total effect accounted for by the mediation of the neurotic subcluster action on CA through lipids was calculated as: β_M_/β_C_ × 100%, where β_C_ is the effect value of the neuroticism subcluster action on CA in univariate MR.

### Statistical Analysis

Data computations and statistical evaluations were conducted using R programming (https://www.r-project.org/, version 4.2.2). MR analyses were performed using the 2SMR package ^50^. Cochran’s Q test was applied with the leave-one-out test to assess the robustness and reliability of the outcomes. The MR-Egger intercept approach was used to assess the genetic pleiotropy. Steiger’s directionality test with the 2SMR package was used to determine the direction of causality. To perform the MR analysis of outcome exposure, the evaluation metrics used were the odds ratio (OR) and 95% confidence interval (95%CI). All statistical *P*-values were two-sided tests, and SNP loci identified using the GWAS study were considered statistically significant at *P* < 5 × 10^-6^; other statistical tests were considered statistically significant at *P* < 0.05.

## RESULTS

### Technology Roadmap

#### Screening of IVs

SNPs exhibiting LD were excluded during the screening for IVs in this study. Thereafter, SNPs associated with the neuroticism subclusters (*ebi-a-GCST006478, ebi-a-GCST006475, ebi-a-GCST006950*) were incorporated, following their alignment with GWAS data pertaining to CA (*ukb-d-I9_CORATHER*). Table 1 presents the screening outcomes for the IVs relevant to each indicator, emphasizing only those that yielded significant results. The *F* statistics for the IVs linked to this indicator exceeded the threshold of 10, indicating that the selected SNPs were predominantly robust IVs, thereby mitigating the potential bias stemming from weak IVs.

**Table 1.**
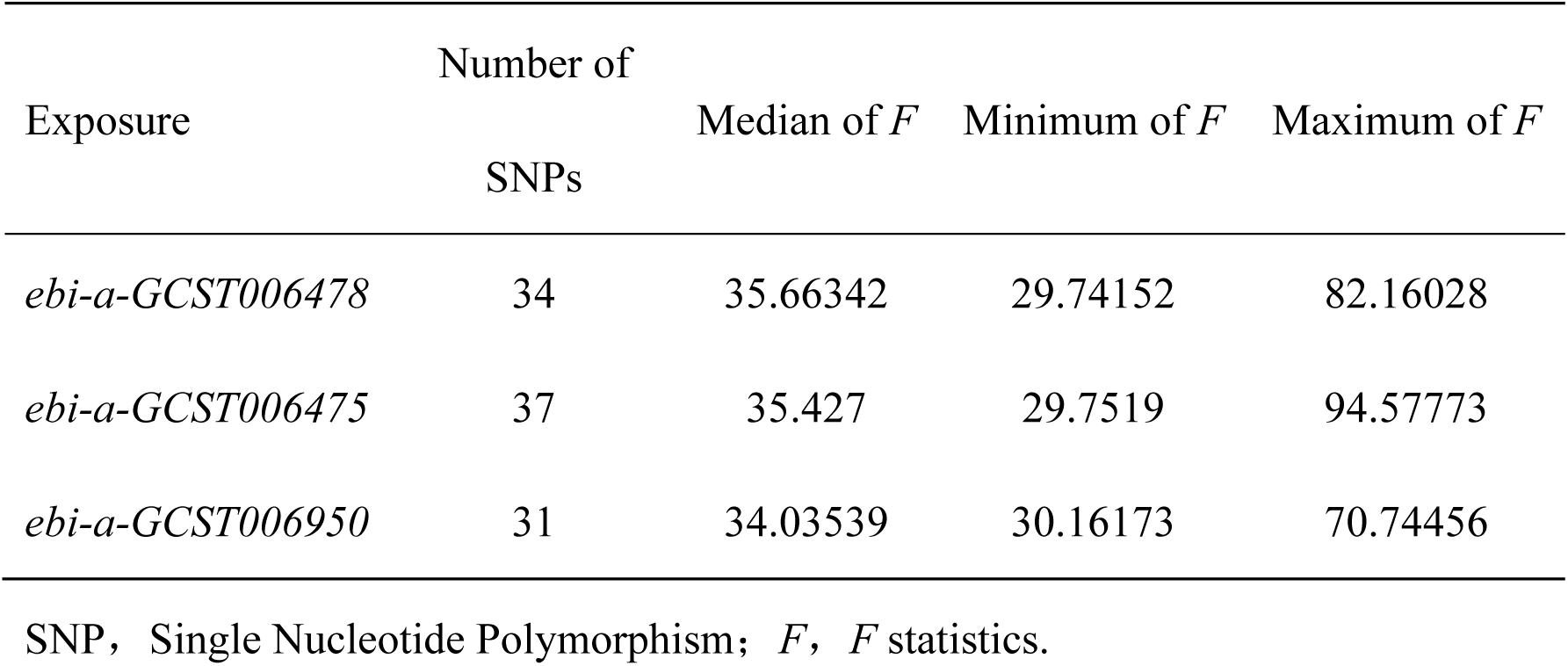
Screening of Instrumental Variables for Neuroticism Subclusters and Coronary Atherosclerosis and *F* Test for the Strength of Instrumental Variables.

| Exposure | Number of<br>SNPs | Median of $F$ | Minimum of $F$ | Maximum of $F$ |
| --- | --- | --- | --- | --- |
| *ebi-a-GCST006478* | 34 | 35.66342 | 29.74152 | 82.16028 |
| *ebi-a-GCST006475* | 37 | 35.427 | 29.7519 | 94.57773 |
| *ebi-a-GCST006950* | 31 | 34.03539 | 30.16173 | 70.74456 |

SNP, Single Nucleotide Polymorphism; $F$ , $F$ statistics.

### Estimation of MR Causal Effects

Five models—MR-Egger, WM, IVW, and simple and weighted modes—were employed to estimate the causal effects of MR. The IVW model revealed a significant causal association between the three neuroticism subclusters (*ebi-a-GCST006478, ebi-a-GCST006475,* and *ebi-a-GCST006950*) and coronary artery disease (*ukb-d-I9_CORATHER*) (Table 2). Furthermore, significant causal relationships were identified between the Worry (*ebi-a-GCST006478*) and CA (OR: 1.01, 95%CI: 1.00–1.02, *P* = 0.026), Depressed affect (*ebi-a-GCST006475*) and CA (OR: 1.02, 95%CI: 1.01–1.03, *P* = 1.52E-05), and Feeling worry (*ebi-a-GCST006950*) and CA (OR: 1.01, 95%CI: 1.00–1.02, *P* =0.036). The compiled results of the MR analysis of the neuroticism subclusters and CA are visually represented in a forest plot (Fig. 2).

**Figure 2.**
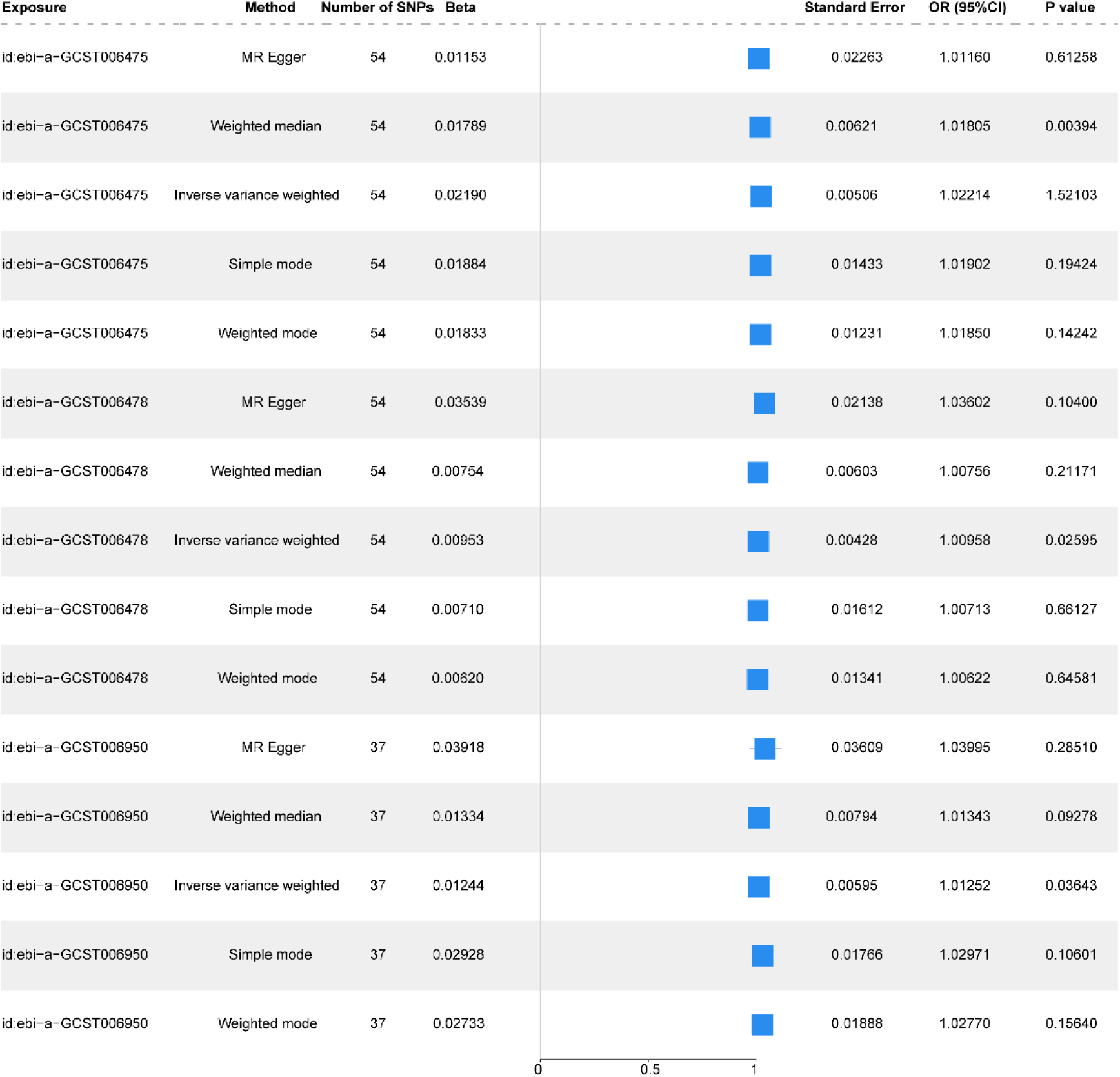
Mendelian Randomization Analysis of the Causal Association between Neuroticism Subclusters and Coronary Atherosclerosis. Forest plot showing the results of the MR analysis of the causal relationship between the two neuroticism subclusters and CA. SNP: single nucleotide polymorphism; OR: odds ratio; CI: confidence interval; MR: Mendelian randomization; CA: coronary atherosclerosis

**Table 2.**
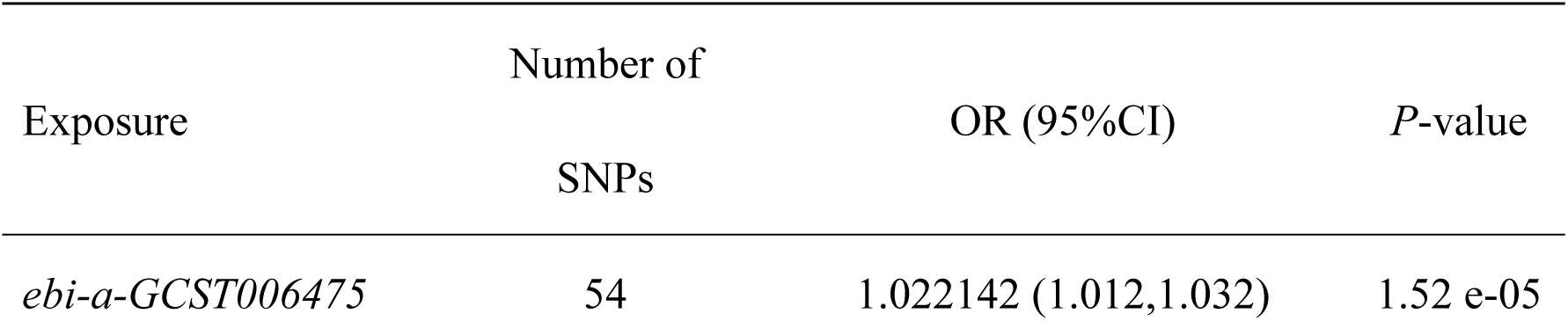
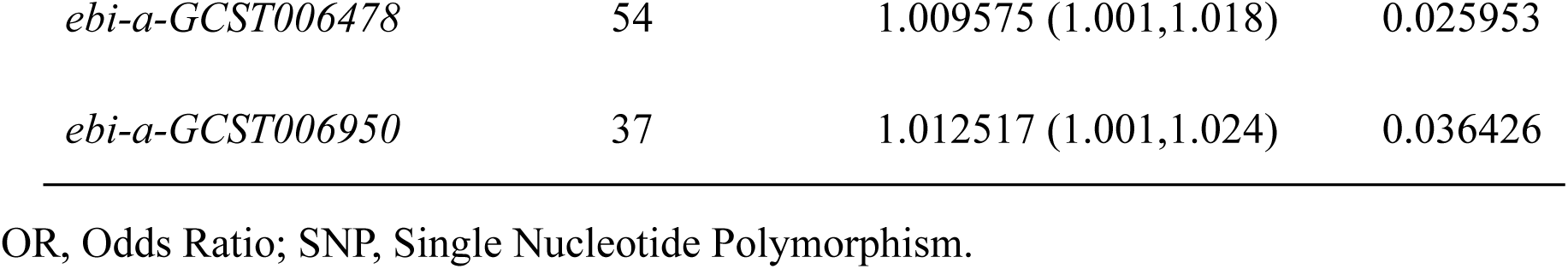
Results of Mendelian Randomization Analysis Assessing the Effect of Neuroticism Subclusters on the Incidence of Coronary Atherosclerosis.

Moreover, Figure 3A–C depicts the linear relationship between the effects of neuroticism subclusters on IVs and their subsequent impact on coronary artery disease incidence across the five analytical models. Therefore, neuroticism subclusters exert a significant influence on CA risk. The linear association observed between the effects of neuroticism subclusters on IVs and their subsequent influence on CA remained consistent across all five analytical models. Specifically, the effect estimates derived from the MR-Egger, WM, simple mode, and weighted mode considerably aligned with results obtained using the IVW model. Furthermore, the slope of these models signifies the magnitude of the β-value, which reflected the strength of the linear relationship between the IVs and the exposure in comparison to the outcome.

**Figure 3.**
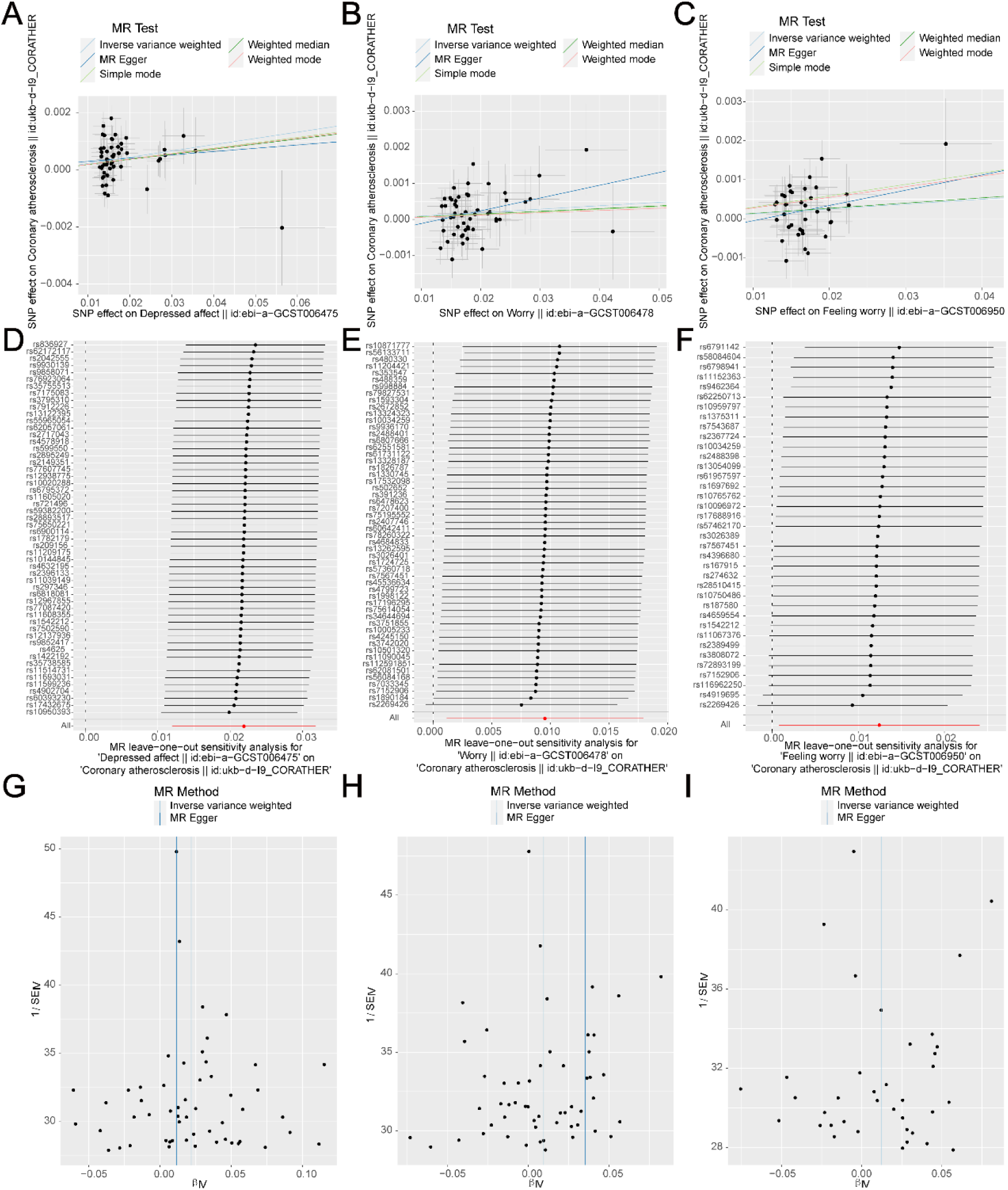
Mendelian Randomization of the Causal Relationship between Neuroticism Subclusters and Coronary Atherosclerosis. A–C. Scatter plots of the results of MR analysis of the causal relationship between two neuroticism subclusters and CA show that the slope of the line indicates the magnitude of the causal relationship predicted by different models. D–F. Forest plots showing the results of the MR analysis of the causal relationship between the two neuroticism subclusters and CA. G–I. Funnel graphs showing the results of the MR analysis of the causal relationship between the two neuroticism subclusters and CA. SNP: single nucleotide polymorphism; CA: coronary atherosclerosis; MR: Mendelian randomization.

We then utilized Cochran’s Q test and the *I²* statistic to evaluate heterogeneity within the results derived from the MR-Egger and IVW models. Notably, no discernible heterogeneity in the MR results concerning CA was observed for the neuroticism subclusters *ebi-a-GCST006478* and *ebi-a-GCST006950* (Cochran’s Q *P* > 0.05). In contrast, MR results for CA for the subcluster *ebi-a-GCST006475* demonstrated moderate heterogeneity (Cochran’s Q *P* < 0.05, *I²* = 28%) (Table S1). We further conducted leave-one-out tests, systematically excluding each IV, to assess its influence on the neuroticism subclusters in relation to the causal effect on CA (Fig. 3D–F). No substantial changes were observed in the overall effects of the IV set. The funnel plot (Fig. 3G–I) corresponding to the IVs of the neuroticism subclusters illustrated a nearly symmetrical distribution of causal association effects, indicating no potential bias in the results.

We conducted an MR-Egger regression analysis to evaluate the pleiotropy within the IVs. The hypothesis test for the intercept term associated with the neuroticism subclusters yielded a *P*-value exceeding 0.05, accompanied by an intercept value close to zero, indicating minimal influence of pleiotropy on the causal inferences drawn from our study (Table 3).

**Table 3.** Tests of Pleiotropy of the Mendelian Randomization Analysis Assessing the Effect of Neuroticism Subclusters on the Onset of Coronary Atherosclerosis.

| Exposure | MR-Egger | Standard Error | *P*-value |
| --- | --- | --- | --- |
| *ebi-a-GCST006475* | 0.000172 | 4.00 e-04 | 0.64 |
| *ebi-a-GCST006478* | 0.00047 | 4.00 e-04 | 0.2228 |
| *ebi-a-GCST006950* | 0.00045 | 6.00 e-04 | 0.4575 |

MR, Mendelian Randomization.

Finally, we assessed the causal directionality of the neuroticism subclusters to CA using the Steiger directionality test (Table 4). This test computes the variance accounted for by SNPs for both exposure and outcome independently (r²). The SNPs derived from our selected metrics explained a greater variance in exposure than in outcome, indicating significant directionality (*P* < 0.05).

**Table 4.**
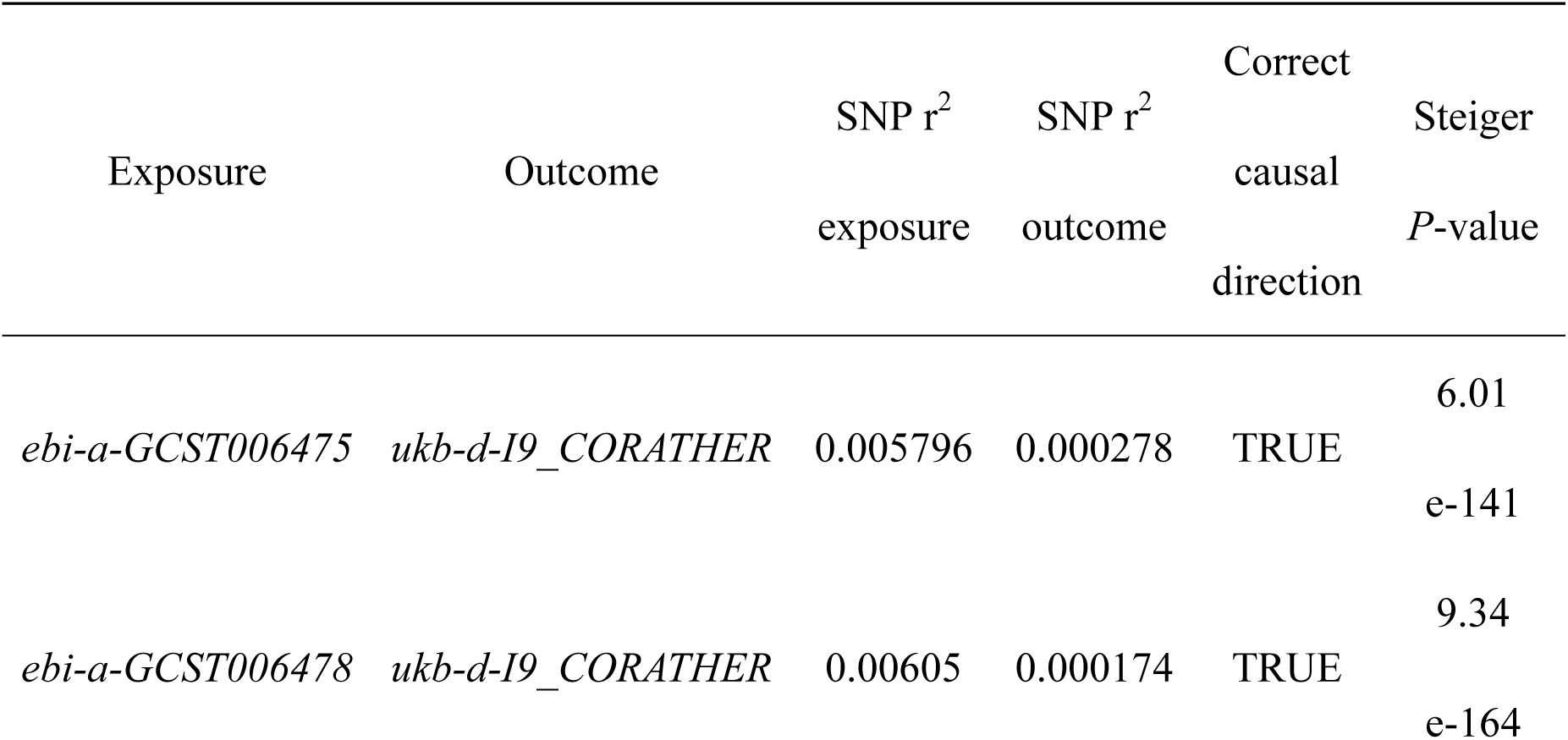
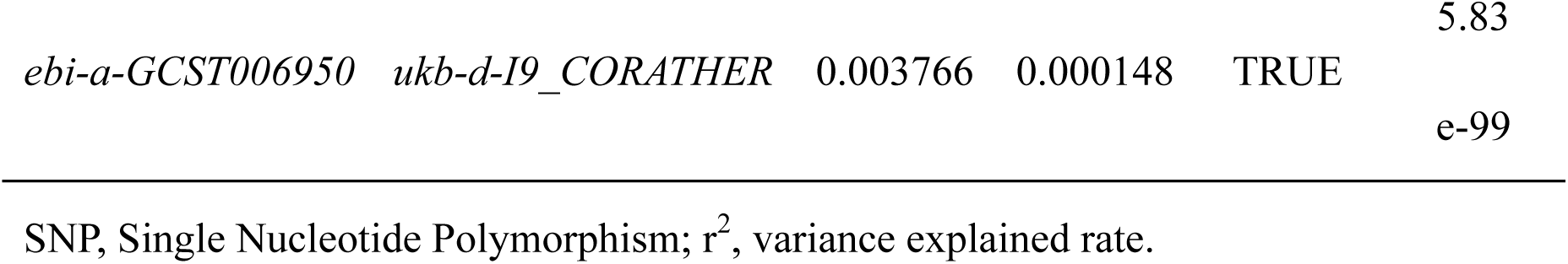
Steiger Directionality Test of the Mendelian Randomization Analysis Assessing the Relationship between Neuroticism Subclusters and Coronary Atherosclerosis.

### Reverse MR Analysis

Among the five analyzed models, the IVW results did not reveal a significant causal relationship between CA and the neuroticism subclusters (*P* > 0.05), indicating that CA is unlikely to influence the neuroticism subclusters. The outcomes of the MR analysis concerning CA and the three neuroticism subclusters were compiled and presented as a forest plot (Fig. 4).

**Figure 4.**
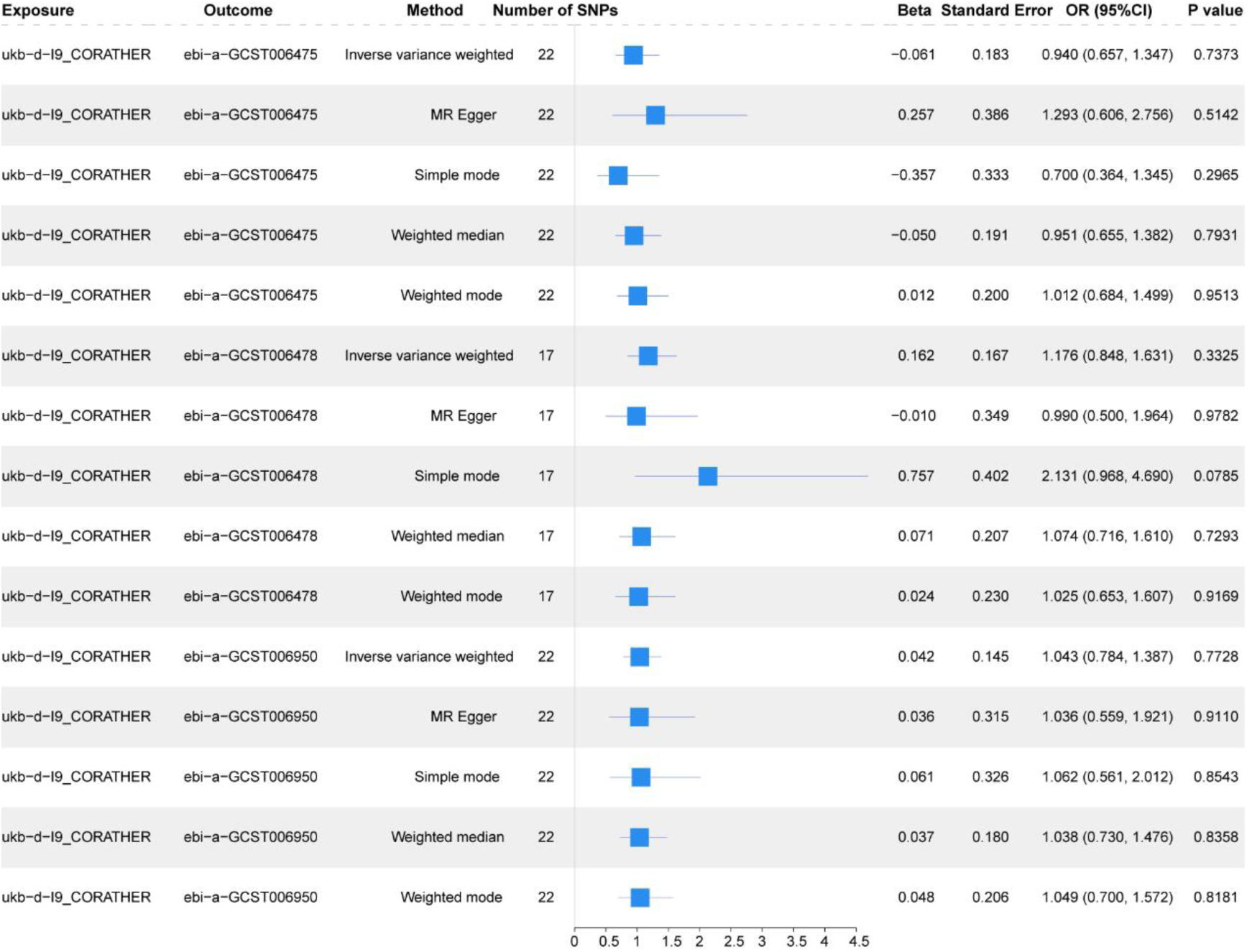
Mendelian Randomization Analysis of the Causal Relationship between Coronary Atherosclerosis and Neuroticism Subclusters. Forest plot showing the results of the MR analysis of the causal relationship between CA and the three neuroticism subclusters. SNP: Single nucleotide polymorphism; OR: odds ratio; CI: confidence interval; MR: Mendelian randomization

### Colocalization Analysis

The colocalization analyses were performed between CA (*ukb-d-I9_CORATHER*) and three neuroticism subclusters (*ebi-a-GCST006478, ebi-a-GCST006475,* and *ebi-a-GCST006950*). SNP (*rs1333042*), which exhibited the lowest *P*-value for its association with CA (*ukb-d-I9_CORATHER*), was designated as the lead SNP. Additionally, we retrieved all SNPs located within a 50 kb radius both upstream and downstream of the lead SNP for colocalization analyses (Fig. 5A, B). The colocalization analyses revealed that CA (*ukb-d-I9_CORATHER*) and the neuroticism subcluster (*ebi-a-GCST006950*) shared a posterior probability of 0.165% (H4 = 0.165%) for the causal variant, whereas the probability for the causal variant between CA (*ukb-d-I9_CORATHER*) and the neuroticism subcluster (*ebi-a-GCST006478*) was 0.167% (H4 = 0.167%). Therefore, neuroticism subclusters did not exhibit a colocalized causal effect on CA. Importantly, the posterior probability of SNP (*rs1333042*) for CA (*ukb-d-I9_CORATHER*) ranged from 40% to 45%.

**Figure 5.**
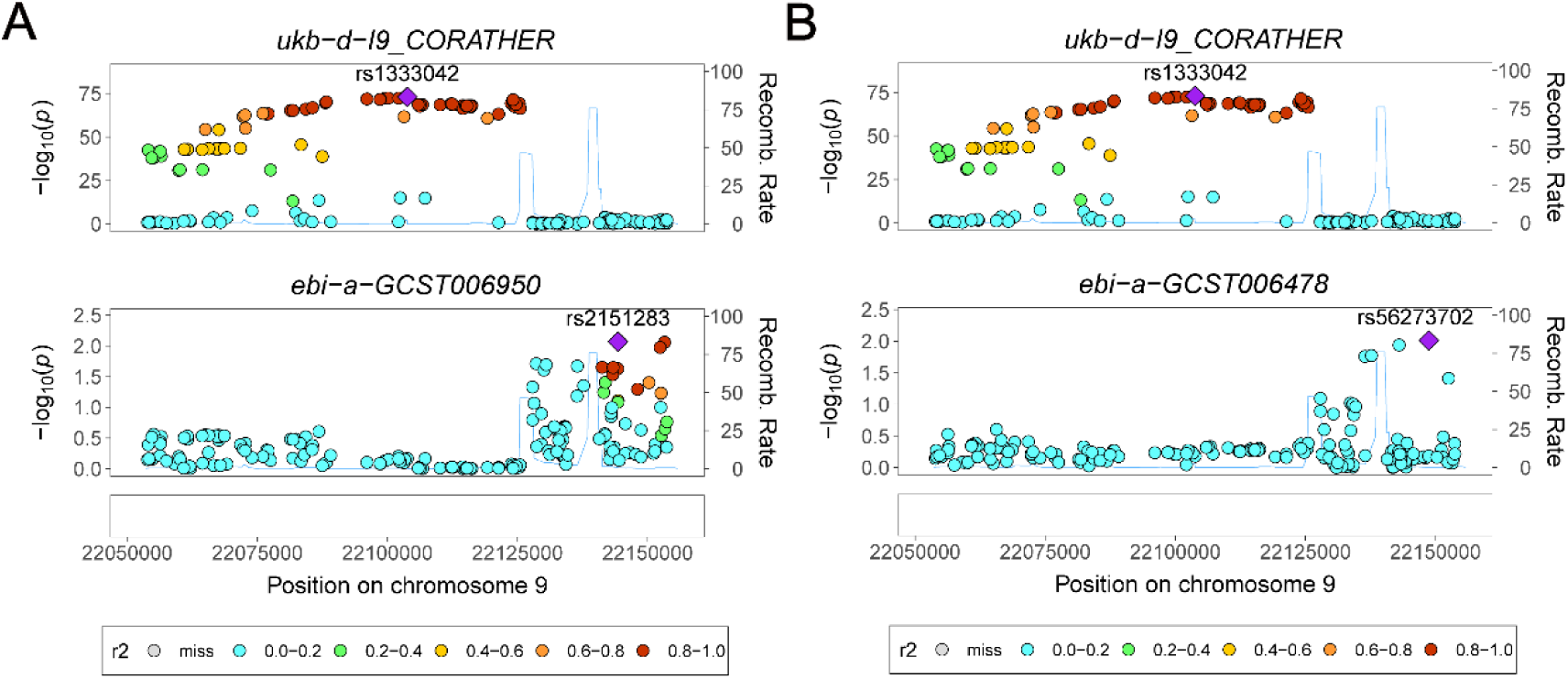
Results of Colocalization Analysis between Coronary Atherosclerosis and Neuroticism Subclusters. A, B. Results of colocalization analysis of coronary atherosclerosis (*ukb-d-I9_CORATHER*) and neuroticism subclusters (*ebi-a-GCST006950* (A), *ebi-a-GCST006478* (B)).

### MVMR Analysis of Neuroticism Subclusters on CA Development

We individually incorporated the risk factors associated with CA, including obesity, diabetes mellitus, and hypertension, and performed MVMR analyses using neuroticism subclusters to assess their direct impacts on CA. We established two by two models that integrated three CA-related risk factors (*finn-bE4_OBESITY*, *ukb-a-306*, and *finn-b-I9_HYPTENS*) along with three neuroticism subclusters (*ebi-a-GCST006478*, *bi-a-GCST006475*, and *ebi-a-GCST006950*) for outcome prediction, resulting in MVMR models (Table 5). The neuroticism subclusters retained a statistically significant association with CA, even after adjusting for the individual influences of obesity, diabetes mellitus, and hypertension (*P* < 0.05).

**Table 5.**
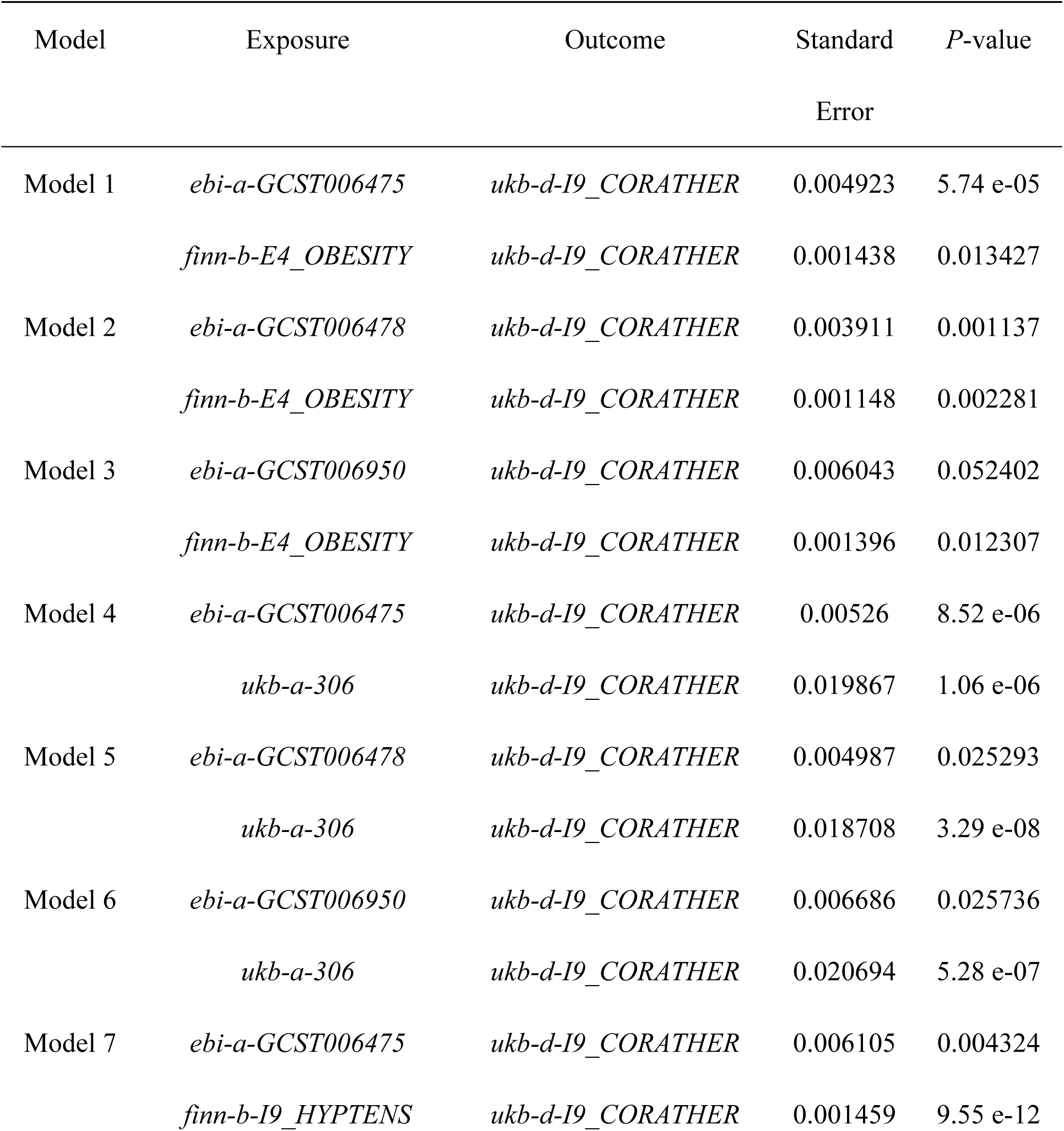
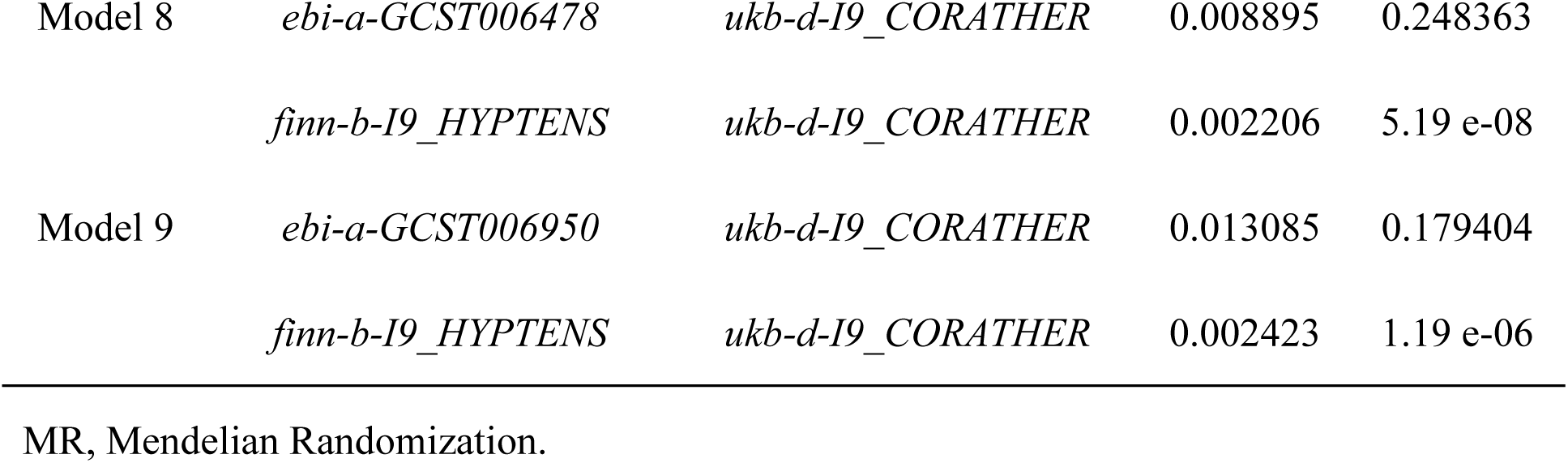
Multivariable Mendelian Randomization Analyses Assessing the Effect of Neuroticism Subclusters on the Development of Coronary Atherosclerosis.

### Analysis of Mediated Effects

We initially applied five models: MR-Egger, WM, IVW, simple mode, and weighted mode. The IVW model revealed a significant causal relationship between *ebi-a-GCST006475* and *ebi-a-GCST006950* and lipids among the screened neuroticism subclusters (Table S2).

Next, the Steiger directionality test was used to determine the accuracy of the causal direction from the neuroticism subclusters to the lipids (Table S3). The Steiger directionality test calculated the variance explained ratio (r^2^) of SNPs for exposure and outcome risk factors. The results demonstrated a significant causal relationship between the selected SNPs and lipid levels for two exposures (*ebi-a-GCST006475* and *ebi-a-GCST006950*). Furthermore, the variance explained was greater for the exposures than for the outcome risk factors in the direction of TRUE with *P* < 0.05, indicating statistical significance.

We then performed an MVMR analysis of neuroticism subclusters together with lipids (Table S4) and assessed the mediating effect of the MVMR analysis, using the model of mediators exhibiting a significant causal relationship with the outcome. The MVMR model revealed a significant effect of lipids on CA (*ukb-d-I9_CORATHER*) (*P* < 0.05), and the neuroticism subclusters (*ebi-a-GCST006475* and *ebi-a-GCST006950*) could potentially affect CA through lipids, constituting a partially mediated effect model.

We calculated the mediating effect values of neuroticism subclusters affecting the development of CA through lipids from the causal effect values of neuroticism subclusters on lipids obtained using univariate MR analyses, as well as the direct and indirect effect values of lipids on CA obtained using MVMR analyses, and the results are presented in Table 6. Notably, neuroticism subclusters (*ebi-a-GCST006475* and *ebi-a-GCST006950*) could affect CA (*ukb-d-I9_CORATHER*) through lipids (Table S5).

**Table 6.**
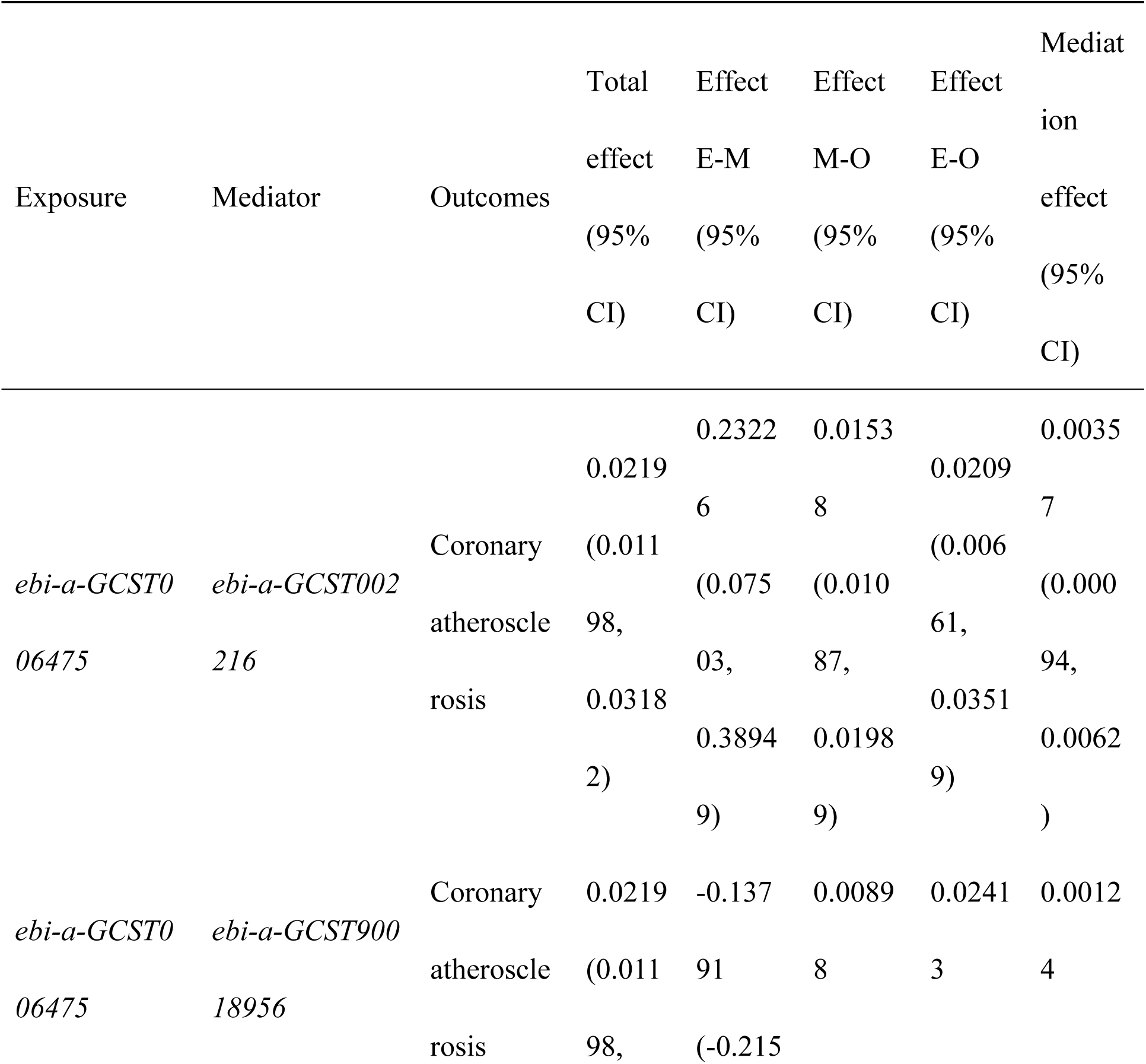
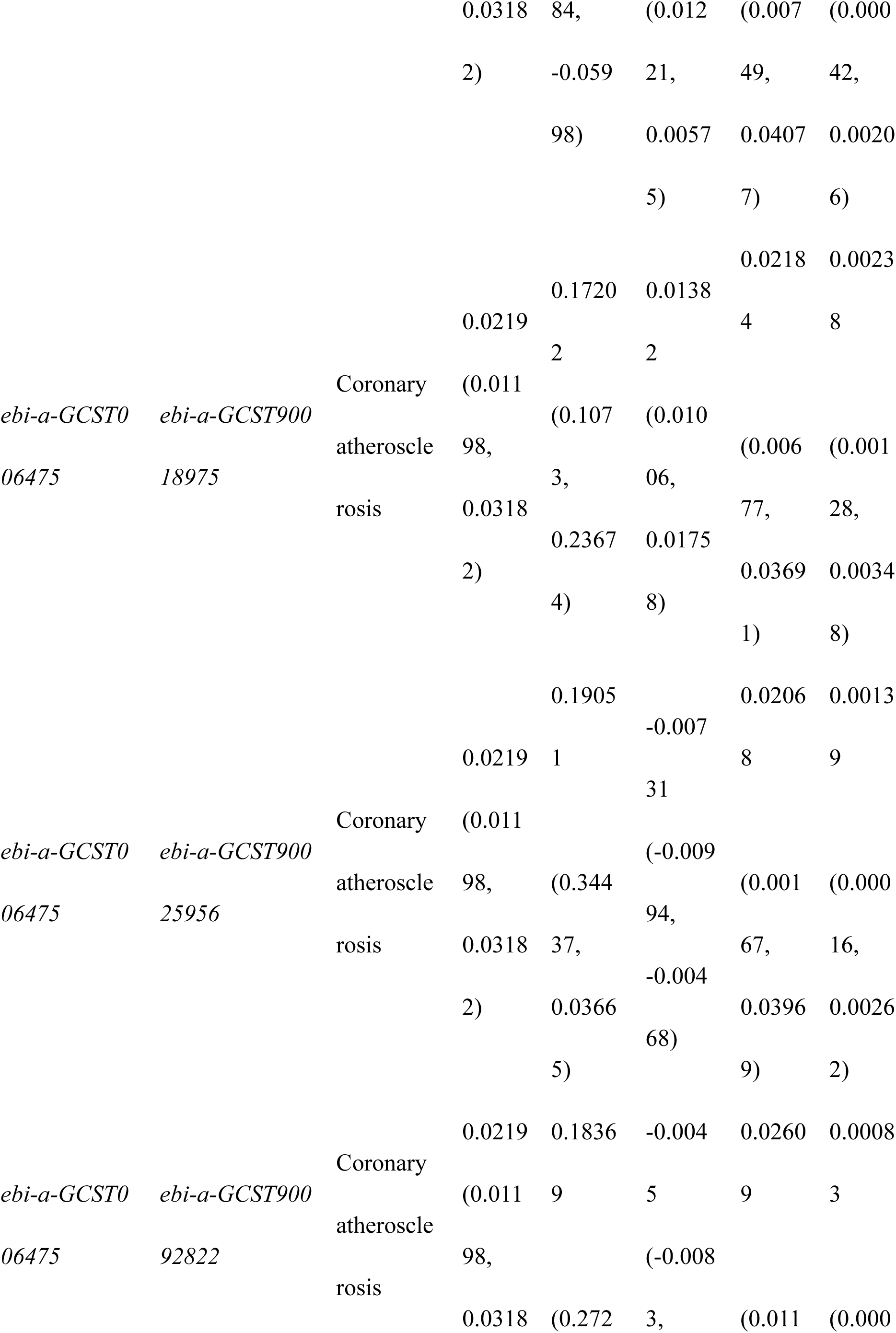
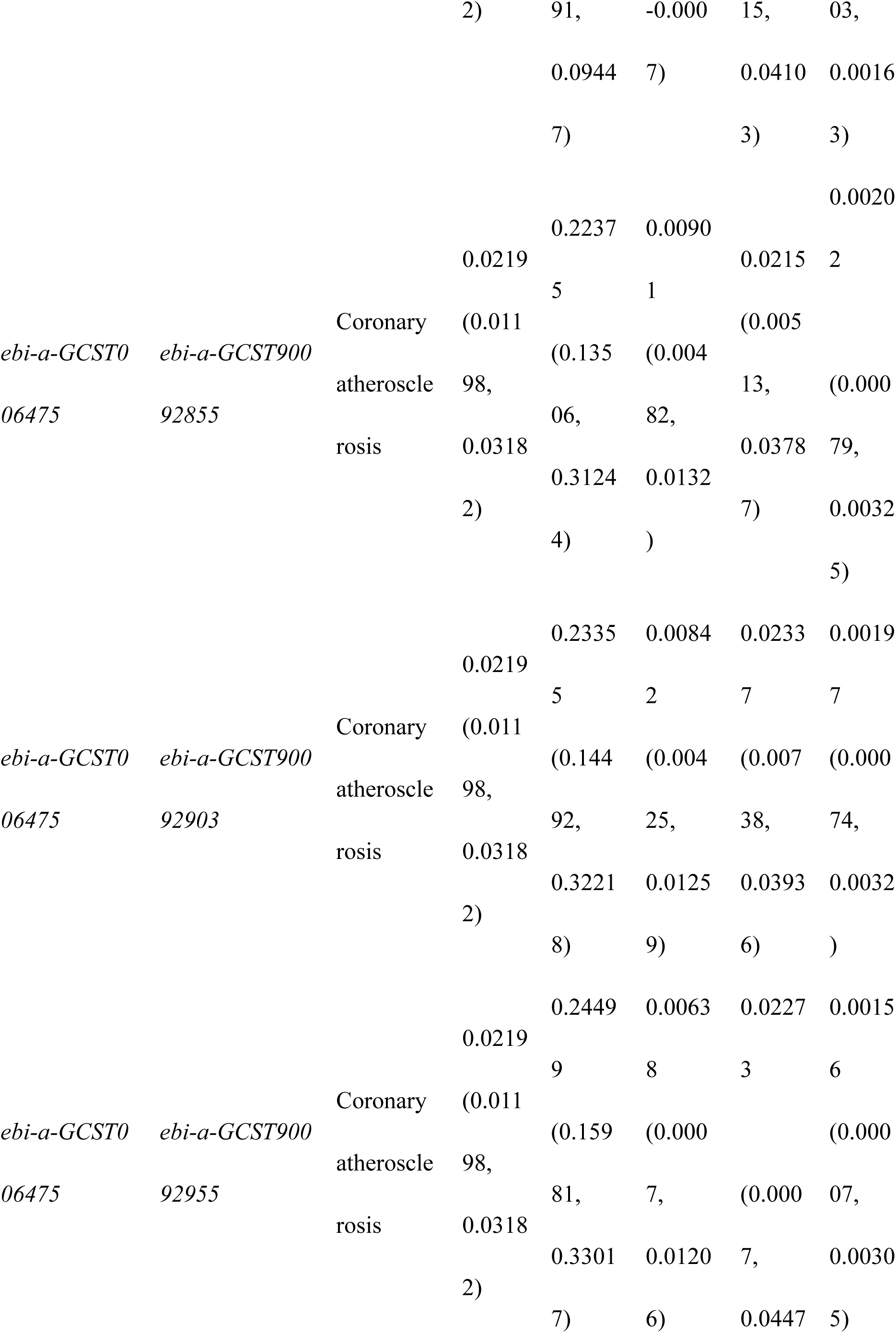
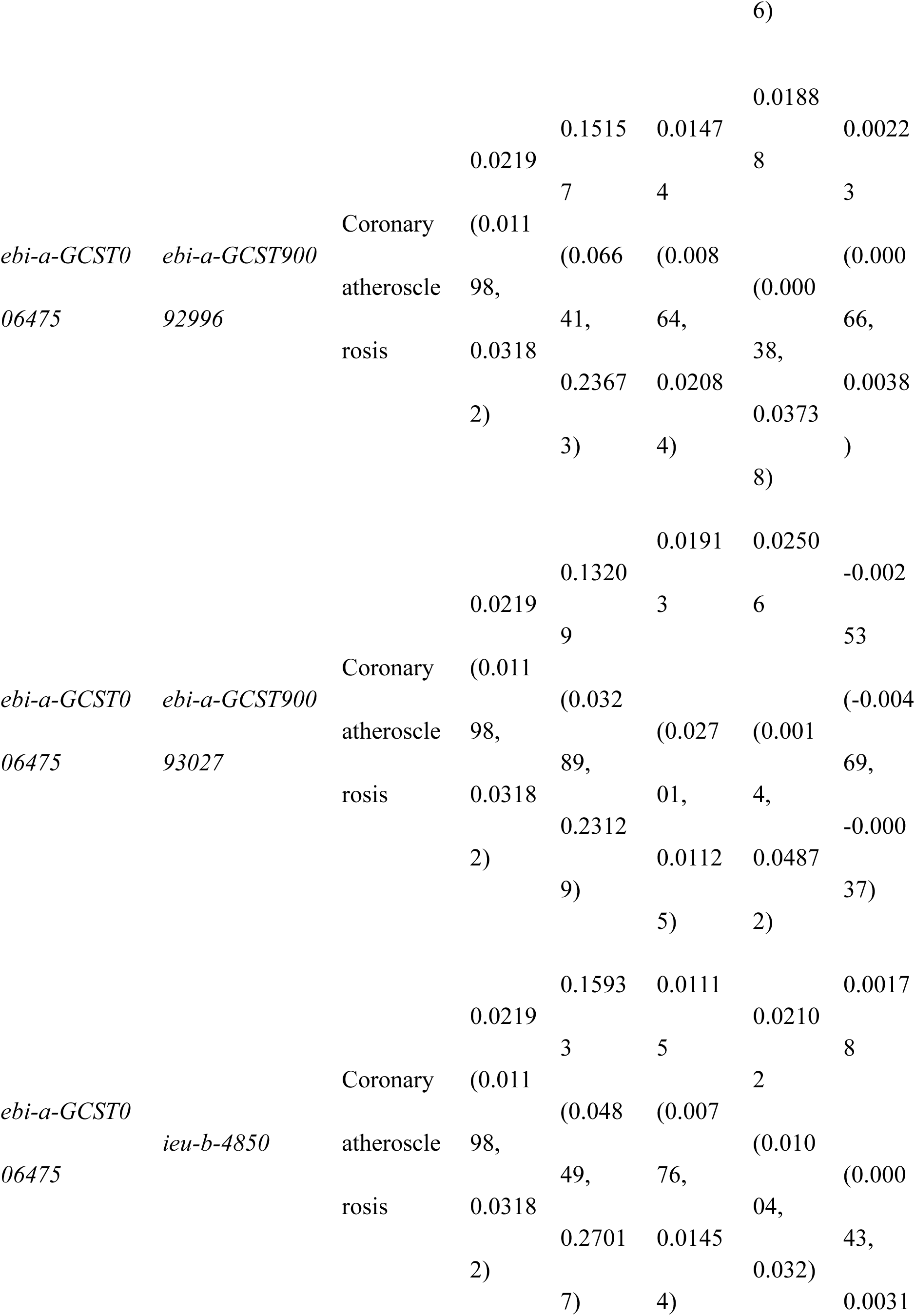
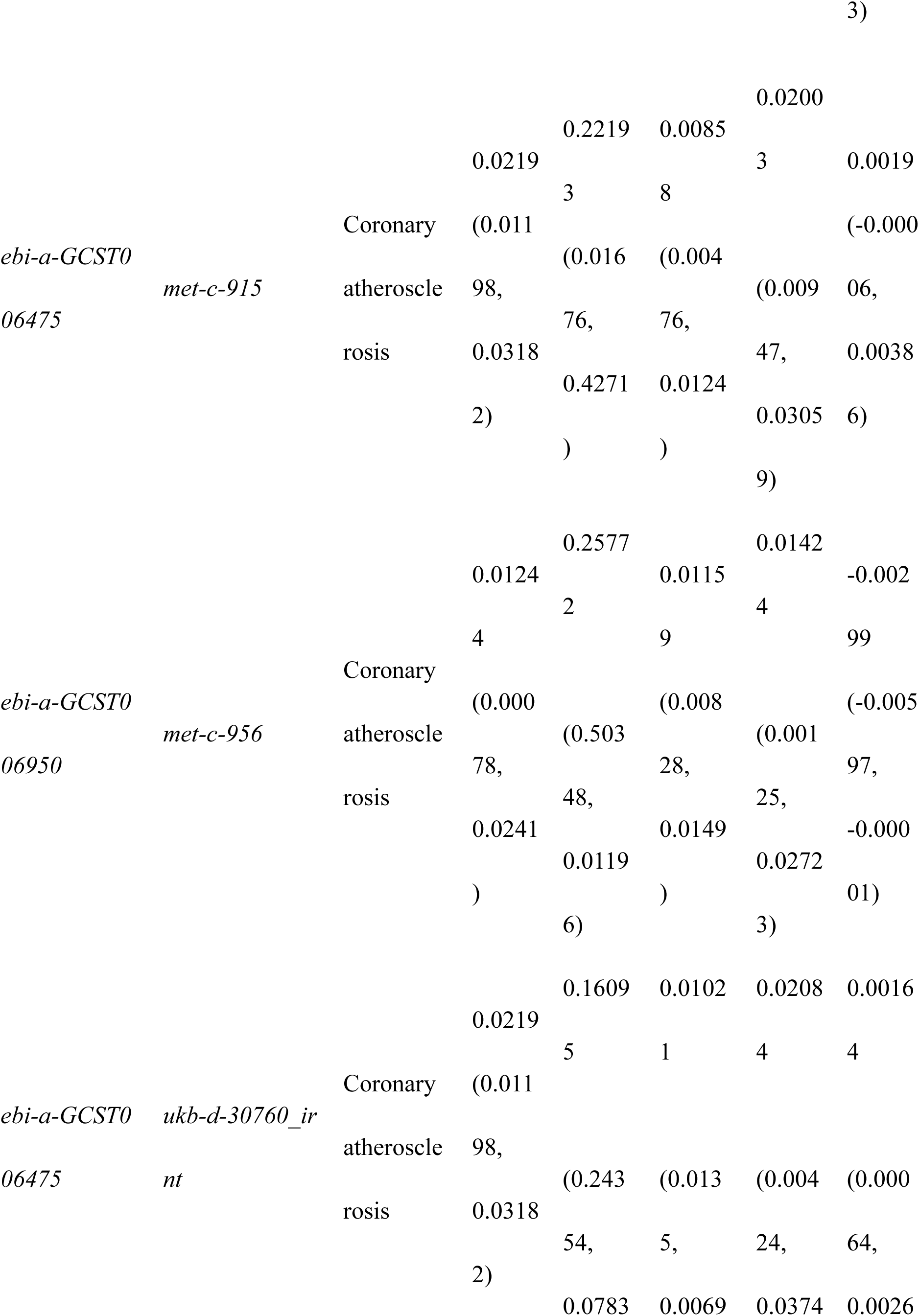
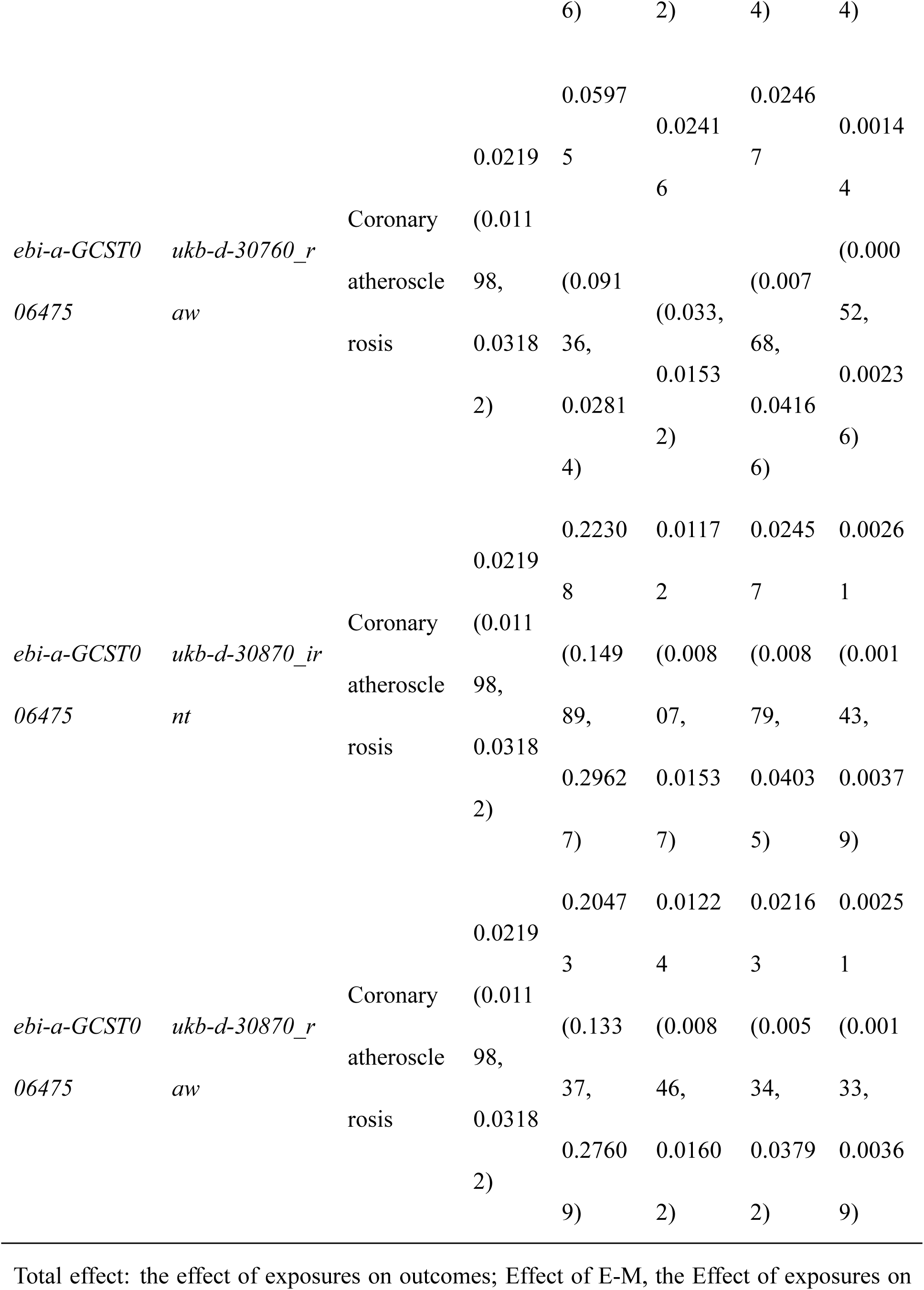
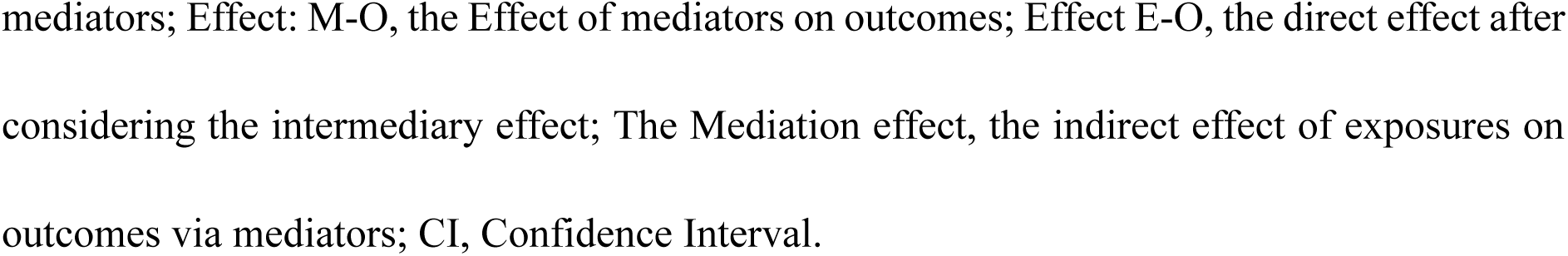
Assessment of the Mediating Role of Lipids in the Causal Relationship Between Neuroticism Subclusters and Coronary Atherosclerosis.

## DISCUSSION

This study examined the relationship between neuroticism subclusters, emphasizing worry and depressive symptoms, and the risk of developing CA using MR analysis to avoid confounders and reverse causation. The primary aim of this study was to elucidate whether these personality traits directly influence CA and the extent to which lipids mediate this relationship. This study utilized the publicly available GWAS dataset for 2SMR and used various MR methods (including MR-Egger, WM, IVW, simple mode, and weighted mode) to estimate causal effects. Sensitivity analyses (heterogeneity, pleiotropy, and leave-one-out tests) were performed to ensure the robustness and reliability of the outcomes. We also performed colocalization analyses to determine whether different phenotypic features were driven by the same causal variants. Our study also employed MVMR to estimate the direct effects of neuroticism subclusters on CA and to calculate lipid-mediated indirect effects. This integrated approach provides a robust framework for understanding the complex interactions between personality traits and CA, potentially offering information for future preventive strategies and therapeutic interventions.

Mood swings can also genetically increase the risk of coronary artery disease, as identified in MR analyses of individual neuroticism programs ^51^. Our findings are consistent with and expand on those of a previous study in this area ^37^. The genetic components associated with neuroticism and CA provide important insights into potential therapeutic targets. Specifically, the identification of SNPs in *ebi-a-GCST006478*, *ebi-a-GCST006475*, and *ebi-a-GCST006950*, which are causally associated with CA, emphasizes the importance of these loci in understanding the interactions between personality traits and cardiovascular health. These genetic markers could be candidate loci for interventions to mitigate the effects of neuroticism on the risk of heart disease in the future. Notably, the effects of genetic factors may vary across populations ^52,53^, warranting further on the variability of these genetic associations across different population groups. Furthermore, gene–environment interactions require exploration, as environmental variables may influence the expression and implications of SNPs on susceptibility to disease ^54,55^. Consequently, these findings in clinical settings might necessitate the formulation of precise prevention strategies, considering both the genetic vulnerability and psychosocial circumstances of vulnerable populations.

In this study, we identified a causal association between CA and the neuroticism subclusters. However, reverse MR analyses revealed no significant causal effect of CA on the neuroticism subclusters, suggesting a unidirectional effect of neuroticism on CA.

Our exploration of pathway interactions revealed a limited degree of evidence for colocalization between neuroticism and CA. However, MVMR analyses demonstrated substantial influence of neuroticism subclusters on CA risk. This association remained significant even when accounting for confounding variables such as obesity, diabetes mellitus, and hypertension, suggesting that neuroticism might independently affect CA susceptibility. Consequently, the relationship between neuroticism subclusters and CA may be multifaceted rather than straightforward. Certain SNPs (e.g., *rs1333042*) exhibited notable posterior probabilities, indicating that although direct genetic overlaps might not be prominently observable, these SNPs could still impact the pathophysiological mechanisms of CA via alternative biological pathways. Future investigations should focus on elucidating these pathways, particularly those associated with inflammation, lipid metabolism, and vascular health ^56–58^, as they might be the crucial mediators of the relationships identified in this study. Moreover, examining how these pathways interact with psychosocial elements may provide valuable insights into integrated intervention strategies that address psychosocial well-being and cardiovascular risk.

Mediation analyses suggested that neuroticism might influence CA risk by modifying lipid profiles. This finding is consistent with that of earlier studies ^59,60^, offering new perspectives on the interplay between psychosomatic factors and metabolic health and indicating that interventions aimed at reducing neuroticism could improve lipid profiles and decrease CA risk ^61^. This further underscores the necessity of enhanced lipid management in individuals exhibiting both neuroticism and CA. Furthermore, investigating whether psychosomatic factors directly influence metabolic markers and how these alterations affect CA prognosis over time is essential. Longitudinal studies are required to substantiate these findings and establish causal relationships. This study serves as a cornerstone of integrated therapeutic approaches involving psychosomatic health strategies.

Incorporating the findings from various MR models into CVD prevention initiatives is essential. The robust and consistent results obtained from these diverse MR methodologies bolster our conclusions regarding identified causal links. Uniformity in both the direction and magnitude of the effects across multiple analytical frameworks reinforces the credibility of our findings. Moreover, this consistency implies that these models may be effectively utilized in broader epidemiological investigations aimed at evaluating causality in complex disease environments ^62^. However, a key issue regarding the criteria that should be used to select the most appropriate model for a given dataset needs to be addressed. Future investigations should prioritize the development of refined modeling techniques encompassing the distinct characteristics inherent in various populations and disease mechanisms. Additionally, novel algorithms designed to enhance the precision of causal inference in genetic epidemiology could substantially advance our understanding of the intricate interplay between genetics, psychiatry, and health outcomes.

In summary, our exploration of the variability in study outcomes revealed that the majority of neuroticism subclusters exhibited a consistent correlation with CA. The moderate heterogeneity detected in specific SNPs highlights the need to consider potential data heterogeneity and interpret the findings with caution. Identifying the origins of this variability is imperative for a precise understanding of our findings and their implications in clinical practice. Factors such as population heterogeneity, environmental influences, and methodological discrepancies may play a role in the observed heterogeneity of outcomes ^63^. Addressing these sources of heterogeneity in future research will enable us to refine our investigation and extend the applicability of our findings across a broader spectrum of contexts. Robust strategies aimed at data harmonization and the integration of cross-study results are vital for progress in this field and for devising more targeted prevention and intervention strategies for at-risk populations.

### Limitations

This study has some limitations that must be acknowledged. First, although we employed MR methodologies to infer causality, the underlying biological mechanisms remain unconfirmed due to the absence of wet laboratory validation. Second, despite the use of multiple datasets, residual batch effects may still exist. Although stringent statistical techniques were used to mitigate these effects, our findings remain confounded. Furthermore, the GWAS data concerning the neuroticism subclusters were calibrated for socioeconomic status, a factor that might have introduced bias into our results. Third, the association between IVs and potential confounders in the 2SMR analysis could not be explored because of the lack of individual-level data. Fourth, our investigation was restricted to participants with European ancestry, limiting the applicability of our conclusions to Asian and African populations. Hence, conducting additional GWAS in diverse ethnic groups is critical to identify a more comprehensive array of SNPs and enhance the quality of MR analyses. Fifth, the absence of long-term follow-up data further constrained our capacity to evaluate the stability and generalizability of our findings across different populations. Sixth, in the mediation analysis, we mainly emphasized the indirect effect of lipids on neuroticism and CA, but other factors, such as heart rate variability ^64,65^ and unhealthy behaviors in the neurotic state ^66^, might play a mediating role, requiring further refinement of our analysis in subsequent studies. Seventh, Cochran’s Q test revealed heterogeneity in certain results, requiring more cautious interpretation. Ultimately, in a two-sample MR context, covariate-adjusted summary associations should generally be avoided, as covariate adjustment may introduce bias even in the absence of pleiotropy ^67^. These limitations suggest that future studies should incorporate experimental validation, long-term follow-up data, and longitudinal research to enhance the robustness and generalizability of our findings.

## CONCLUSION

Our study suggests that neuroticism subclusters are associated with an increased risk of CA and that lipids might mediate this effect to some extent. The risk of CA in neurotic groups should be more carefully assessed in clinical settings, emphasizing lipid management in patients with comorbidities, such as neuroticism and CA. These findings provide new insights into the complex interplay between psychosomatic factors and cardiovascular health and offer potential avenues for future research and therapeutic interventions.

## Data Availability

All data are publically available, so no permission is required.

## ACKNOWLEDGMENTS

We would like to thank the participants and investigators of the database we used in this study. This work was supported by grant funding from the leading Talent Training Plan of Shanghai Pudong New Area Health Commission China (PWRd2022-3), the Key Research and Development Program of Shanghai (ZXS003R2-2-1).

## DISCLOSURES

### Authors’ Contributions

**Jie Ming**: Conceptualization; Data curation; Software; Formal analysis; Investigation; Visualization; Methodology; Writing—original draft; Writing—review and editing. **Rui-lin Li**: Conceptualization; Validation; Investigation; Writing—review and editing. **Yi-li Gao**: Investigation; Methodology. **Shu-ting Huang**: Investigation; Methodology. **Xiao-yan Yao**: Investigation; Methodology. **Qi Tan**: Writing—review and editing. **Ming Zong**: Resources; Funding acquisition; Writing — review and editing. **Lie-ying Fan**: Conceptualization; Supervision; Funding acquisition; Project administration; Writing—review and editing.

### Availability of Data and Materials

All data are publically available, so no permission is required.

### Ethics Approval and Consent to Participate

All GWASs received ethical approval from relevant institutional review boards, informed consent from participants, and rigorous quality control. Since this study is derived from summarized publicly available data, ethical approval was not required for this study.

### Nonstandard Abbreviations and Acronyms

CA: Coronary atherosclerosis
CVD: Cardiovascular disease
GWAS: Genome-wide association studies
TG: Triglycerides
LDL-C: Low-density lipoprotein cholesterol
ApoB: Apolipoprotein B
HDL-C: High-density lipoprotein cholesterol
ApoA: Apolipoprotein A
MR: Mendelian randomization
IV: Instrumental variable
RCT: Randomized controlled trial
SNP: Single nucleotide polymorphism
LD: Linkage disequilibrium
IVW: Inverse variance-weighted
WM: Weighted median
2SMR: Two-sample mendelian randomization
MVMR: Multivariable mendelian randomization

